# Evaluation of Amyloid Removal as a Surrogate for Cognitive Decline: Pilot Analysis in Individual-Level Data from the A4 Study of Solanezumab

**DOI:** 10.1101/2025.07.21.25331942

**Authors:** Sarah F. Ackley, Michael Flanders, Audrey Murchland, Ruijia Chen, Jingxuan Wang, Sachin J. Shah, Edward D. Huey, M. Maria Glymour

## Abstract

**Background:** Amyloid removal has been used as a surrogate outcome in Alzheimer’s disease trials, allowing accelerated approval of aducanumab and lecanemab. The A4 (Alzheimer’s Clinical Trial Consortium A4/LEARN) trial’s individual-level data supports novel methods to evaluate amyloid’s validity as a surrogate for cognitive decline.

**Methods:** In 812 participants, cognitive and functional change was measured using the CDR-SB score. Instrumental-variable analysis estimated the effect of amyloid reduction; mediation analysis quantified solanezumab’s cognitive effect mediated by amyloid reduction.

**Results:** Each 10 centiloid reduction due to randomization to treatment was associated with 0.026 higher CDR-SB (95% CI: -0.013, 0.065) 14.6% of solanezumab’s effect on cognition was mediated by amyloid reduction (95% CI: –122%, 208%).

**Discussion:** Estimates showing near-zero effects of amyloid reduction on cognitive decline suggest minimal impact of amyloid reduction in populations with little disease progression. Replication in anti-amyloid trials with larger treatment effects could guide treatment and regulatory decisions.

## Introduction

Under the Food and Drug Administration’s Accelerated Approval Program, amyloid removal has served as a surrogate marker for cognitive and functional outcomes in Alzheimer’s disease trials. Aducanumab (1) and lecanemab were approved based on amyloid reduction (2) Subsequently, lecanemab and donanemab received full approval by demonstrating cognitive and functional benefits (3). Rigorous evaluation of amyloid removal as a surrogate endpoint for cognitive decline using clinical trial data has not been reported.

While some argue that the trial results for recent agents clearly demonstrate a causal role of amyloid in cognitive and functional decline (4,5), others speculate about the likelihood of non-amyloid mechanisms. Ongoing skepticism about the safety and efficacy of amyloid-targeting drugs persists (6,7). With better tools to analyze and integrate results from existing trials, there is the potential to move beyond opinion-based debates toward evidence-based conclusions grounded in rigorous analysis. Toward this goal, several prior meta-analyses used aggregated, trial-arm level data from trials of anti-amyloid drugs to evaluate amyloid removal’s effects on cognitive decline with conflicting results; the most recent of which finds small overall benefits of amyloid removal (8–12)

However, existing literature that attempts to formally evaluate the effect of amyloid reduction on cognitive decline has significant limitations due to lack of individual-level clinical trial data. Specifically, existing analyses assume drug effects are fully mediated by changes in amyloid. This assumption can be tested in individual-level data, but not in group-level data. In addition, group-level averages may mask important features of amyloid-cognition relationships, such as widely hypothesized non-linear associations (13,14). Finally, additional statistical assumptions may be required when working with aggregated data (8)

Drawing on methods from epidemiology and econometrics (15,16), we propose the following sequential analyses for evaluating surrogate outcomes in individual-level clinical trial data: 1) using causal mediation analysis, quantify the fraction of the drug effect on the outcome that is mediated by the putative surrogate outcome (17); 2) using IV analysis, take the ratio of the effect of randomization on the outcome to the effect of randomization on amyloid reduction, which we interpret as the effect of treatment on the outcome per unit change in the surrogate. If, in analysis 1, the drug effect is largely through the putative surrogate, then the effect of treatment on the outcome per change in the surrogate approximates the causal effect of surrogate on the outcome (15)

Taken together, if these analyses demonstrate that the surrogate mediates a substantial portion of the treatment effect (17) and that changes in the surrogate result in a clinically meaningful improvement in the outcome under realistic conditions, this provides strong evidence for the surrogate’s validity. Typical approaches to evaluate surrogacy examine within-arm correlations between the putative surrogate and outcome (18,19). In contrast, our approach takes advantage of the benefits of randomization in IV analyses to evaluate whether amyloid is an appropriate surrogate for clinical outcomes and has the major advantage of allowing for within-arm adjustment of confounding in mediation analyses.

The full implementation of these proposed methods requires individual-level data on change in amyloid and change in cognition or function over time, from individuals randomized to varying doses of anti-amyloid drugs (including placebo). Their application in this context has been hindered by a lack of familiarity with these methods for evaluation of surrogate outcomes and by restricted access to individual-level data from anti-amyloid drug trials. In this manuscript, we first illustrate our approach via simulated data. We then reanalyze the newly available individual-level Anti-Amyloid Treatment in Asymptomatic Alzheimer’s Disease (A4) study data, a large study of solanezumab for prevention of cognitive decline in preclinical dementia.

Specifically, we demonstrate how these approaches can be used with individual-level data to evaluate the suitability of amyloid as a surrogate for cognitive change in clinical trials. As additional trial data becomes available (e.g, from the TRAILBLAZER-ALZ-2 trial of donanemab (20)), we recommend these approaches be implemented to rigorously assess amyloid reduction as a surrogate outcome for cognitive decline.

## Methods

### A4 Study

The Anti-Amyloid Treatment in Asymptomatic Alzheimer’s Disease (A4) study was a phase 3 randomized-controlled clinical trial of solanezumab, an anti-amyloid monoclonal antibody, for the reduction of cognitive decline in individuals with elevated amyloid assessed via amyloid PET but no clinical symptoms (21). Eligibility criteria required a Clinical Dementia Rating Global Scale (CDR-GS) score of 0, MMSE greater than or equal to 25, and a Wechsler Memory Scale Logical Memory score between 6 and 18 inclusive, and elevated amyloid on florbetapir-PET. Apolipoprotein E (APOE) genotyping was required for all study participants. Enrollment began in 2014 and 1169 participants aged 65-86 were randomized to receive intravenous solanezumab or placebo. Participants were followed for at least 240 weeks (nearly 5 years), with cognitive assessments performed regularly and neuroimaging performed at the start and end of the study (240 weeks) and into the open-label extension. A treatment dose escalation was approved mid-trial, with dosages increasing from 400 to 1200 mg. Additional details on changes to the treatment protocol and COVID-19-related visit timing disruptions are reported elsewhere (21–24). The study ended in 2022, and the final results reported in 2023 indicated that solanezumab did not slow cognitive decline (25)

We perform secondary data analysis using de-identified individual-level data from the A4 Study. All participants provided informed consent at the time of enrollment. Institutional review board (IRB) approval was obtained at each of the trial sites. The use of deidentified data in this analysis was determined to be exempt from additional IRB review via consultation with the Brown University IRB staff. Data were obtained from the Anti-Amyloid Treatment in Asymptomatic Alzheimer’s Disease (A4) and Longitudinal Evaluation of Amyloid Risk and Neurodegeneration (LEARN) Study Data Package. Data is available to qualified researchers with a short application at a4studydata.org

### Amyloid-PET Imaging

As described elsewhere (22), all participants underwent amyloid Positron Emission Tomography (PET) imaging. Imaging outcomes are reported for the first and last study visits. PET images were acquired 50 to 70 minutes after receiving an injection of florbetapir. Amyloid burden was quantified using a mean cortical standardized uptake value ratio (SUVr) calculated based on a whole cerebellar reference region. Amyloid-PET SUVrs were converted to Centiloids using the following formula: Centiloids = 183.07*(Florbetapir SUVr) – 177.26.

### Cognitive and Functional Outcomes

Full details on cognitive testing are described elsewhere (26). Change in the Preclinical Alzheimer Cognitive Composite (PACC) score at 4.5 years was the A4 study’s primary endpoint. For broad interpretability, we use a selection of the PACC subcomponents which contain the following global and domain-specific cognitive measures: mini-mental state examination (MMSE) and digit symbol substitution test (DSST, Wechsler adult intelligence scale). The clinical dementia rating sum of box (CDR-SB) cognitive and functional assessment was also performed regularly, as well as the Alzheimer’s Disease Cooperative Study–Activities of Daily Living (ADCS-ADL) functional assessment. We focus on results for the CDR-SB since it is the primary outcome in more recent and successful trials of monoclonal antibody drugs targeting amyloid. The CDR-SB is the only assessment for which a higher score represents worse function. Additional information on the range, domain(s) evaluated, and associated dementia-related cutoffs for cognitive outcomes in the PACC is given in Supplemental **Table S1.** We refer to all cognitive and functional outcomes evaluated as cognitive outcomes for clarity and parsimony with some loss of linguistic precision.

### Analysis

All analyses were performed using the R statistical computing environment (version 4.4.1). Only complete-case analyses were performed.

#### Estimates of Change and Covariates

Change in amyloid for each participant was determined by taking the difference between Centiloids collected at baseline and the final study visit at approximately 240 weeks. We estimated cognitive change using an annual trajectory by taking the slope of a linear model fit with respect to time, giving cognitive change per year for each individual. This procedure allows for the use of participants with missing follow-up assessments and accommodates differences in study visit timing due to the COVID-19 pandemic. Sensitivity analyses are performed using change in CDR-SB from the first visit where CDR-SB was assessed to the 5-year visit, and using change in composite PACC (Preclinical Alzheimer’s Cognitive Composite) score from the first visit to the 5-year visit. Pearson correlations are calculated for treatment arm (1 for solanezumab and 0 for placebo) and amyloid change, and number of days from baseline to dosage escalation and amyloid change.

Analyses were adjusted for sex, APOE-ε4 gene dose (0, 1, or 2 copies), baseline age, baseline cognition, and baseline amyloid. Since participants were randomly assigned to treatment, adjustment for baseline cognition would not be expected to induce bias (27). Descriptives were calculated for covariates and estimates of change. Relationships between cognitive and functional score change and amyloid change were plotted.

#### Mediation and Instrumental-Variable Analyses

##### Description of Methods for Evaluation of a Surrogate

**Figure 1** provides conceptual models (directed acyclic graphs) of mediation and instrumental variable (IV) analyses for examining the effect of amyloid removal. Mediation analysis and IV analysis are complementary methods in this context (28)

**Figure 1:**
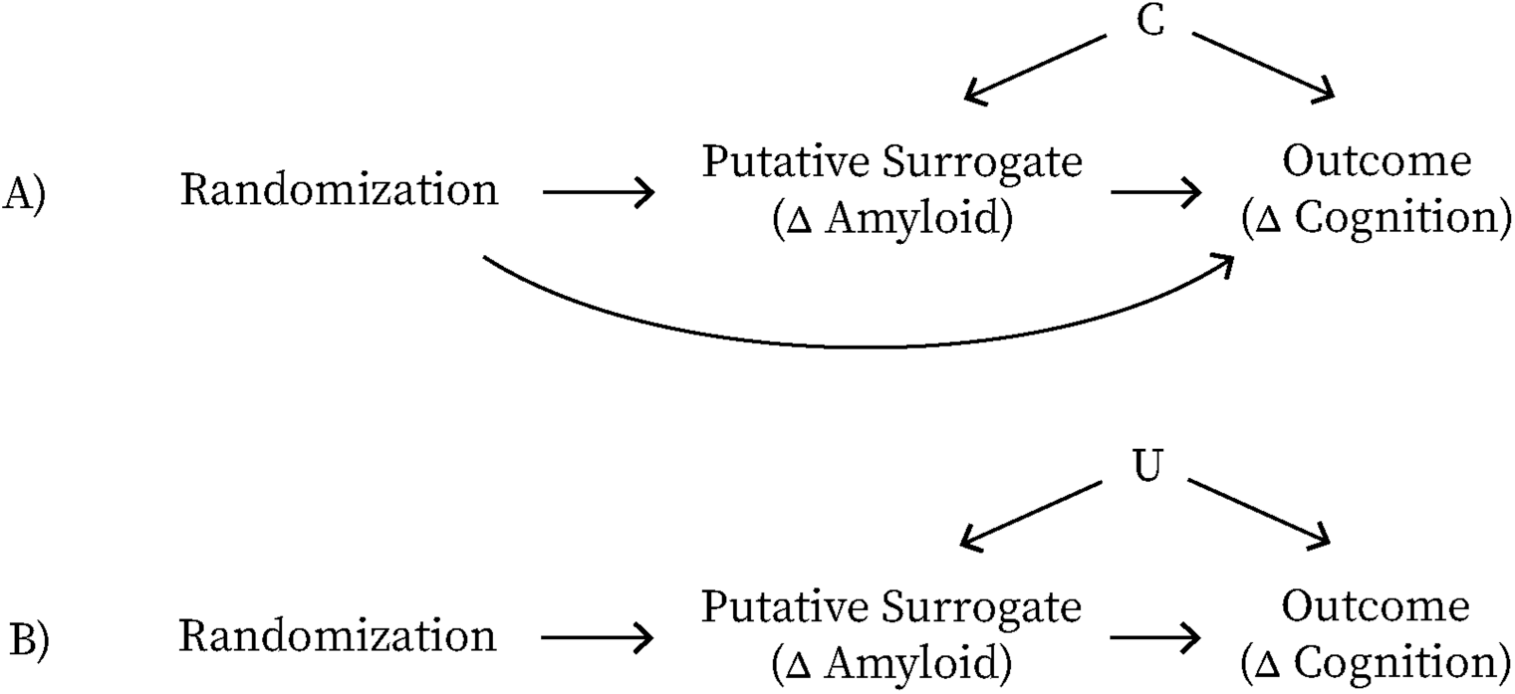
Conceptual models of instrumental-variable and mediation analyses for analyses examining the effect of amyloid removal. (A) The effect of anti-amyloid therapy on cognition is partially mediated by change in amyloid, but confounders C (e.g., sex, APOE-ε4, and baseline age, cognition and amyloid) influence both change in amyloid and change in the outcome. Mediation analyses estimate the fraction of the effect of randomization on cognition that operates via changes in amyloid, but requires that we know all common causes (confounders) C of change in amyloid and change in cognition. (B) The instrumental variable (IV) estimate provides the ratio of the effect of randomization to treatment on cognition to the effect of randomization on change in amyloid; under the stronger assumption that the drug’s effect operates entirely via amyloid, this provides an estimate of the effect of amyloid removal on functional and cognitive change. Because this assumption may not be met, we interpret the IV estimate as the effect of randomization to treatment on cognition, scaled by the change in amyloid.

**Mediation analysis** was developed across multiple disciplines and is used to determine the extent to which a causal effect is explained by a specific mechanism, i.e., operates via hypothesized mediating variables (29–31) *Causal* mediation methods developed by epidemiologists are considered the field standard now, since they address problems with previous mediation methods, specifically they allow for accurate estimation of direct and indirect effects even in the presence of exposure-mediator interaction and flexibly accommodate non-linear effects(30). These and similar methods have been applied in the context of evaluating surrogate outcomes previously (17,32)

Similar to within-arm analyses, causal mediation requires that we know and adjust for all common causes of the mediator and outcome, here confounders of amyloid change and cognitive change. For drugs that rapidly reduce amyloid, the assumption of minimal unmeasured confounding may be reasonable. Rapid amyloid clearance induced by these treatments likely overwhelms natural variability in accrual, minimizing the influence of endogenous factors that affect amyloid accrual and cognitive decline. On the other hand, with the strategy of near-total amyloid removal with donanemab in TRAILBLAZER-ALZ 2, baseline levels effectively represent the total amyloid removed, suggesting that any determinants of total amyloid at baseline could act as confounders. In our analyses, we adjust for potential predictors of total amyloid and rate of amyloid accrual, such as APOE-ε4 and age.

**Instrumental variable** (IV) methods were largely developed in econometrics and address unmeasured confounding by leveraging a randomized source of variation that influences a mediator (33). Random assignment is referred to as an instrument for the effect of the mediator on the outcome (34)^1^ if the effect of randomization on the outcome is entirely via the mediator, i.e., there are no direct effects of the instrument on the outcome. In this setting, we can calculate an IV estimate that corresponds to the causal effect of the mediator on the outcome – in our setting, the causal effect of amyloid change on cognitive change. To obtain a valid estimate of causal effect, there is no need to adjust for or even know what common causes the mediator and outcome share; one need only assume the mediator is the only possible mechanism linking randomization to the outcome.

Adherence-adjusted effect estimates are a common application of IV estimates in the medical literature, because they can provide unconfounded estimates of the effect of perfect adherence to a treatment on an outcome (34). This is a strength when compared to per-protocol and as-treated estimates, which are biased if there is confounding of the treatment and outcome. As argued previously, an IV approach can be applied to randomized controlled trials to scale intention-to-treat effect estimates by changes in a biomarker that is a putative surrogate (15) Our approach in this work is analogous to adherence-adjusted effects, but here leverages randomization to produce an unconfounded estimate of the biomarker’s effect on the outcome (15)

In cases where the drug effects are not fully mediated by the surrogate, the IV estimate does not provide the causal effect of the surrogate on the outcome. In such cases we describe the IV estimate as the randomization effect on the outcome scaled per unit change in the surrogate.

This is useful in the context of both formal meta-analysis and in informally comparing drugs across trials since it expresses estimates of different dosages of different drugs on the same scale, allowing direct comparisons. In addition to improving comparability, such estimates in and of themselves can be used to evaluate whether the effect is in fact through the surrogate. Specifically, we can derive IV estimates of the effect of randomization on cognition per unit reduction in amyloid from multiple trials of different drugs. If these IV estimates are not approximately equal, this suggests that amyloid-targeting drugs may not be operating entirely via changes in amyloid (15). Alternatively, inconsistencies in IV estimates could represent differences in factors like population, disease stage, or rates of progression across trials.

##### Simulation of Illustrative Data

To illustrate how mediation and IV estimates depend on relationships between change in cognition and change in amyloid, we simulated data to visualize the relationships under three scenarios. Scenarios X and Y were based on observed changes in amyloid from the CLARITY-AD and TRAILBLAZER-ALZ-2 trials of lecanemab and donanemab (20,35). Scenario Z has a much smaller difference in change in amyloid between groups as observed in the A4 trial. For all scenarios, data was simulated using correlated normal and skewed-normal distributions representing amyloid and cognitive change, respectively. For scenarios X and Y, the treatment group was simulated with greater reductions in amyloid – though perhaps not as great as that observed in recent trials (mean = -70, sd = 30). The placebo group was simulated with slight increases in amyloid (mean = 15, sd = 30). For scenario Z, both the treatment and placebo arms had slight increases in amyloid (mean = 15, sd = 30). For all scenarios, a correlation (ρ = 0.35) was assumed between amyloid change and cognitive decline to model the hypothesis that greater reductions in amyloid are associated with slower cognitive decline. Cognitive change was generated from a skew-normal distribution with a skew parameter of 5 to reflect observed asymmetry in cognitive decline trajectories. We modeled significant cognitive decline (mean = 1.4, sd = 0.5) in the placebo arm for scenario X, and in both the placebo and drug arms for scenario Y. In scenario X, we optimistically assume treatment halts cognitive decline (mean = 0, sd = 0.5), while in scenario Y, treatment has no effect on cognition (mean 1.4, sd = 0.5). In scenario Z, we model no cognitive change in both placebo and drug arms (mean = 0, sd = 0.5) .

##### Implementation

We conducted causal mediation analyses to estimate the extent to which changes in amyloid mediate the cognitive effects of randomization to solanezumab treatment. We specified a series of linear regression models to evaluate the relationships between treatment assignment, amyloid change, and cognitive outcomes. The first model, the mediator model, regressed amyloid change on treatment group assignment to estimate the effect of solanezumab versus placebo on amyloid change. The second model, the outcome model, regressed cognitive change on both amyloid change and treatment group, as well as confounders for adjusted models. For outcome models, we tested for an interaction between randomization and change in amyloid in producing cognitive change; interaction terms were not included since none were significant (p-interaction > 0.1). The proportion of the total effect mediated through amyloid change is calculated as the ratio of the mediated effect (average causal mediation effect) to the total effect (average treatment effect) using the mediate function from the R mediation package (version 4.5.0) with the two models as inputs. Confidence intervals were obtained using quasi-Bayesian Monte Carlo with 1000 simulations (36,37) IV methods were used to estimate the causal effect of amyloid reduction on cognitive changes using randomization as an instrument for amyloid reduction. Similar to mediation, IV requires separate models to estimate the relationships between the randomization instrument, the mediator, and the outcome. However, R-based implementations of IV regression integrate these steps into a single estimation step. Specifically, we estimated the effect of randomization on cognitive change per change in amyloid using the ivreg function from the R AER package (version 1.2.14). In IV analyses, we adjust for the same confounders as in the mediation analyses in both regressions, but this is for the purpose of improving precision and not for confounding adjustment. IV estimates are scaled to present estimates as the effect of treatment on annual cognitive change per 10 Centiloid *reduction* in amyloid. A 10 Centiloid reduction is used because 10 Centiloids is roughly the difference in change in Centiloids between treatment arms in A4. In addition, using a 10-Centiloid reduction facilitates rescaling results to produce expected cognitive effects for trials of drugs that more effectively remove amyloid and are comparable to prior IV estimates (9)

##### Assessment of Non-Linearity

To assess non-linearities using IV, multiple instruments, such as multiple treatment arms or other arbitrary sources of variation in dose, are needed. This is because the relationship between change in amyloid and change in cognition is assessed by treatment arm. During the A4 trial, a dose escalation was introduced to increase amyloid clearance. The timing of this escalation varied across participants, depending on when they enrolled in the study, effectively introducing a form of pseudorandomization. Participants who underwent dose escalation earlier in the study would be expected to be exposed to a higher cumulative dose of solanezumab, likely resulting in greater amyloid reduction.

We used the visit number of dose escalation as an instrumental variable to assess non-linear effects of amyloid reduction on cognitive outcomes using an F-test for an amyloid-squared term. This approach to assess nonlinearity using the visit number of treatment escalation will not be generalizable to other contexts. For trials with three or more treatment arms, nonlinearity can be evaluated using the IV methods we present. However, for trials with only two treatment arms, assessing nonlinearity will either require pooling data from multiple studies (or interacting randomization with another variable, requiring additional assumptions). This necessity underscores the importance of making trial data publicly accessible to enable evaluations of drug mechanisms.

## Results

For 812 study participants with complete pre/post amyloid-PET measures, complete CDR-SB measures, and at least 2 observations for all other cognitive measures evaluated, the mean (standard deviation [SD]) age was 71.47 (SD 4.45) years and 59.2% of participants were women. The between-group difference (treatment minus placebo) in average amyloid change was -8.32, 95% CI (−11.17, -5.30) Centiloids. Average between-group difference in annual change in CDR-SB was 0.02, 95% CI (−0.01, 0.06) points per year. The correlation between treatment arm and amyloid change was -0.19, 95% CI (−0.26,-0.12), p<0.01. Among the solanezumab treated (N=396), correlation between dose escalation day relative to baseline and amyloid change was -0.07, 95% CI (−0.17,0.02), p=0.17. Summary statistics for additional quantities are given in **Table 1**

**Table 1:** Characteristics of treated and untreated groups and overall. Mean and standard deviation are shown for all covariates used in the analysis, along with mean changes in function, cognition, and amyloid. Abbreviations: *CDR-SB: clinical dementia rating sum of box score; MMSE: Mini-Mental State Examination; DSST: Digit Symbol Substitution Test (Wechsler Adult Intelligence Scale); ADCS-ADL: Alzheimer’s Disease Cooperative Study–Activities of Daily Living. PACC: Preclinical Alzheimer’s Cognitive Composite*.

|  | Placebo<br>(N=416) | Solanezumab<br>(N=396) | Overall<br>(N=812) |
| --- | --- | --- | --- |
| **Age** | 71.48 (4.46) | 71.46 (4.43) | 71.47 (4.45) |
| **Sex** |  |  |  |
| Female | 247 (59.4%) | 234 (59.1%) | 481 (59.2%) |
| Male | 169 (40.6%) | 162 (40.9%) | 331 (40.8%) |
| **APOE-ε4 Allele Copies** |  |  |  |
| 0 | 161 (38.7%) | 150 (37.9%) | 311 (38.3%) |
| 1 | 222 (53.4%) | 210 (53.0%) | 432 (53.2%) |
| 2 | 33 (7.9%) | 36 (9.1%) | 69 (8.5%) |
| **Baseline Amyloid Burden on PET, centiloids** | 65.81 (31.05) | 65.60 (31.46) | 65.71 (31.23) |
| **Pre/Post Change in Amyloid PET, centiloids** | 21.12 (20.63) | 12.80 (21.74) | 17.06 (21.57) |
| **Baseline CDR-SB Score** | 0.05 (0.17) | 0.06 (0.16) | 0.05 (0.17) |
| **Change in CDR-SB Score per Year** | 0.10 (0.22) | 0.13 (0.27) | 0.11 (0.25) |
| **Baseline MMSE Score** | 28.79 (1.24) | 28.86 (1.21) | 28.83 (1.22) |
| **Change in MMSE Score per Year** | -0.09 (0.36) | -0.10 (0.41) | -0.09 (0.39) |
| **Baseline DSST Score** | 49.36 (9.61) | 49.05 (10.31) | 49.21 (9.95) |
| **Change in DSST Score per Year** | -0.41 (1.31) | -0.41 (1.38) | -0.41 (1.34) |
| **Baseline ADL Partner Score** | 43.48 (2.66) | 43.56 (2.38) | 43.52 (2.53) |
| **Change in ADL Partner Score per Year** | -0.30 (0.97) | -0.40 (1.12) | -0.35 (1.04) |
| **Baseline PACC Score** | 0.11 (2.59) | 0.21 (2.71) | 0.16 (2.65) |
| **Change in PACC Score per Year** | -0.22 (0.77) | -0.25 (0.91) | -0.23 (0.84) |

Figure 2 is intended to facilitate the interpretation of the mediation analysis results for this and future studies. The results of the mediation analysis are needed for the interpretation of the IV estimate, which is why we present mediation results first in text and figures. Specifically, if amyloid mediates 100% of the effect of solanezumab treatment on cognition, the IV estimate corresponds to the causal effect of amyloid on cognition. Otherwise, the IV estimate gives the effect of randomization to treatment on cognition, scaled per 10 Centiloids of amyloid change; this estimate may over- or under-estimate the effect of amyloid reduction on cognitive change, depending on the percent mediated.

**Figure 2:**
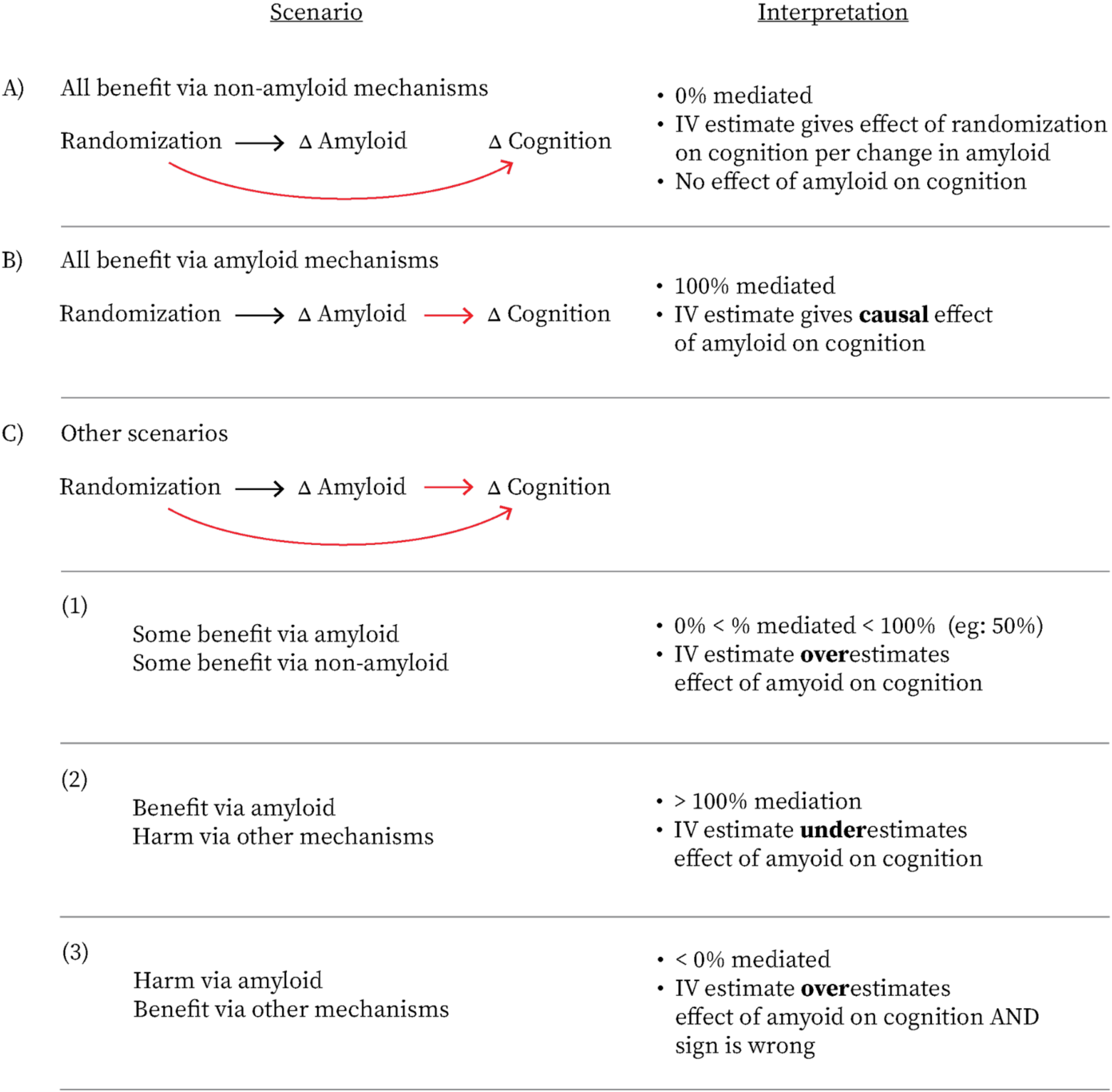
Guide to Evaluation of Amyloid Removal as Surrogate Outcome Using Mediation and IV Analyses. Idealized scenarios in which change in amyloid has no effect on cognition (A) or in which randomization effects cognition only via change in amyloid (B). More realistic (41) scenarios (C1, C2, and C3) assume that randomization influences cognition via multiple mechanisms, including amyloid reduction. In this setting, the interpretation of the estimate is guided by the mediation estimate. Here, we assume an IV estimate that is positive reflecting the putative assumption that amyloid removal reduces cognitive decline. If a cognitive or functional measure, such as the CDR-SB, where increases reflect cognitive worsening is used, or surrogate causes harm, the same reasoning applies to the sign-reversed quantity. Confounders of the relationship between amyloid reduction and functional and cognitive decline are omitted for parsimony, but must be adjusted for to obtain valid mediation results; adjustment for such confounders is not essential for IV estimates but may improve precision.

Figure 3 shows the distribution of changes in the clinical outcome (cognitive function) and potential surrogate (amyloid) for simulated scenarios X, Y, and Z and for individuals in the A4 study. In scenario X, a drug effectively removes amyloid and reduces cognitive decline. In scenario Y, a drug effectively removes amyloid but does not reduce cognitive decline. Finally, in scenario Z, a drug has minimal effect on amyloid and is tested in a population where the placebo group does not clinically progress. In the A4 trial, placebo and treatment groups are similar with very little change in amyloid, cognition, or function. **Figure S1** shows group mean and median scores for all visits for each cognitive and functional measure used. **Figure S2** shows changes in these scores during the trial period using either a modeled trajectory, as in primary analyses, or using difference between first and last visit, as in sensitivity analysis.

**Figure 3:**
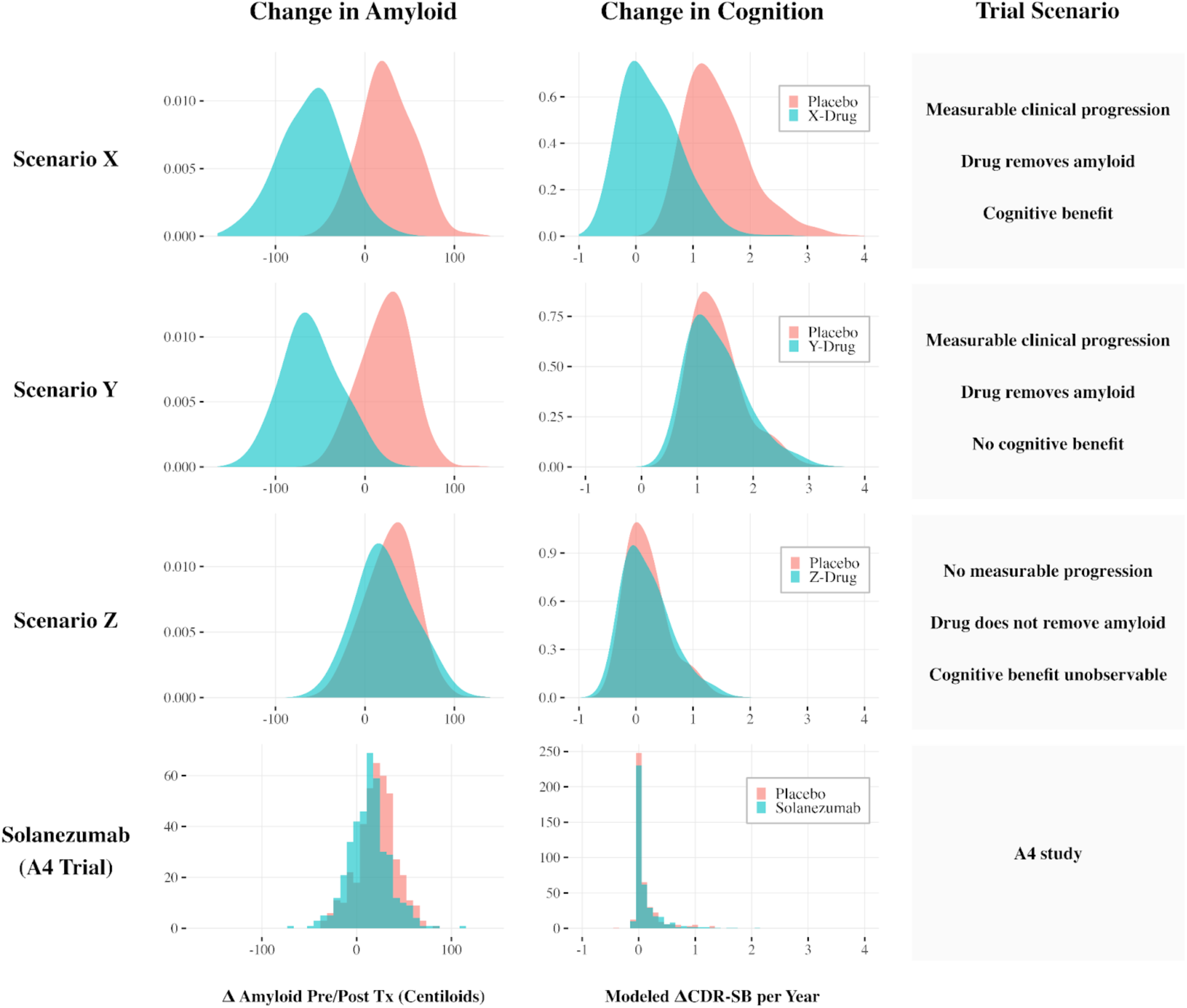
Change in amyloid and change in cognition for simulation scenarios X, Y, and Z and as observed in the A4 trial. Simulated scenarios X and Y are based on mean and standard deviation change in amyloid data from CLARITY-AD and TRAILBLAZER-ALZ 2. In these scenarios, treatment effectively removes amyloid. In scenario X, treatment confers significant functional and cognitive benefit, while treatment confers no functional and cognitive benefit in scenario Y. In scenario Z, we altered the scenario Y simulation, such that there is only a small difference between amyloid change in treated and placebo groups and no average disease progression in either group. In A4, there is a small difference in amyloid change between the placebo and Solanezumab groups and neither functioning nor cognition changed for most participants in the treated or placebo arms.

Figure 4 shows change in amyloid and change cognition and function for individuals, as well as mediation and IV estimates, for hypothetical trials X, Y, and Z, and for the A4 study. Mediation analysis suggests that amyloid change mediates 14.6% of solanezumab’s effect on cognitive change, 95% CI: (−122%, 208%). The IV-estimated effect of treatment on annual change in CDR-SB change per 10 Centiloid reduction was 0.026 (95% CI: -0.013, 0.065). Given that solanezumab’s effect on change in CDR-SB per 10 Centiloids is near 0 with narrow confidence intervals, its amyloid reduction would not be expected to meaningfully impact CDR-SB over five years in populations without cognitive decline, irrespective of the proportion mediated by amyloid (since 0 times any number is 0). The remaining IV and mediation results are qualitatively similar and shown in **Table 2**. There was no evidence of non-linear effects of amyloid on CDR-SB by treatment group in a model where group was defined as the visit number of dose escalation (p=0.27) (plotted in **Figure S3**). Sensitivity analyses performed using change in cognitive and functional outcomes from the first in-study assessment visit and the end-of-study visit are shown in **Tables S2** and **S3**

**Figure 4:**
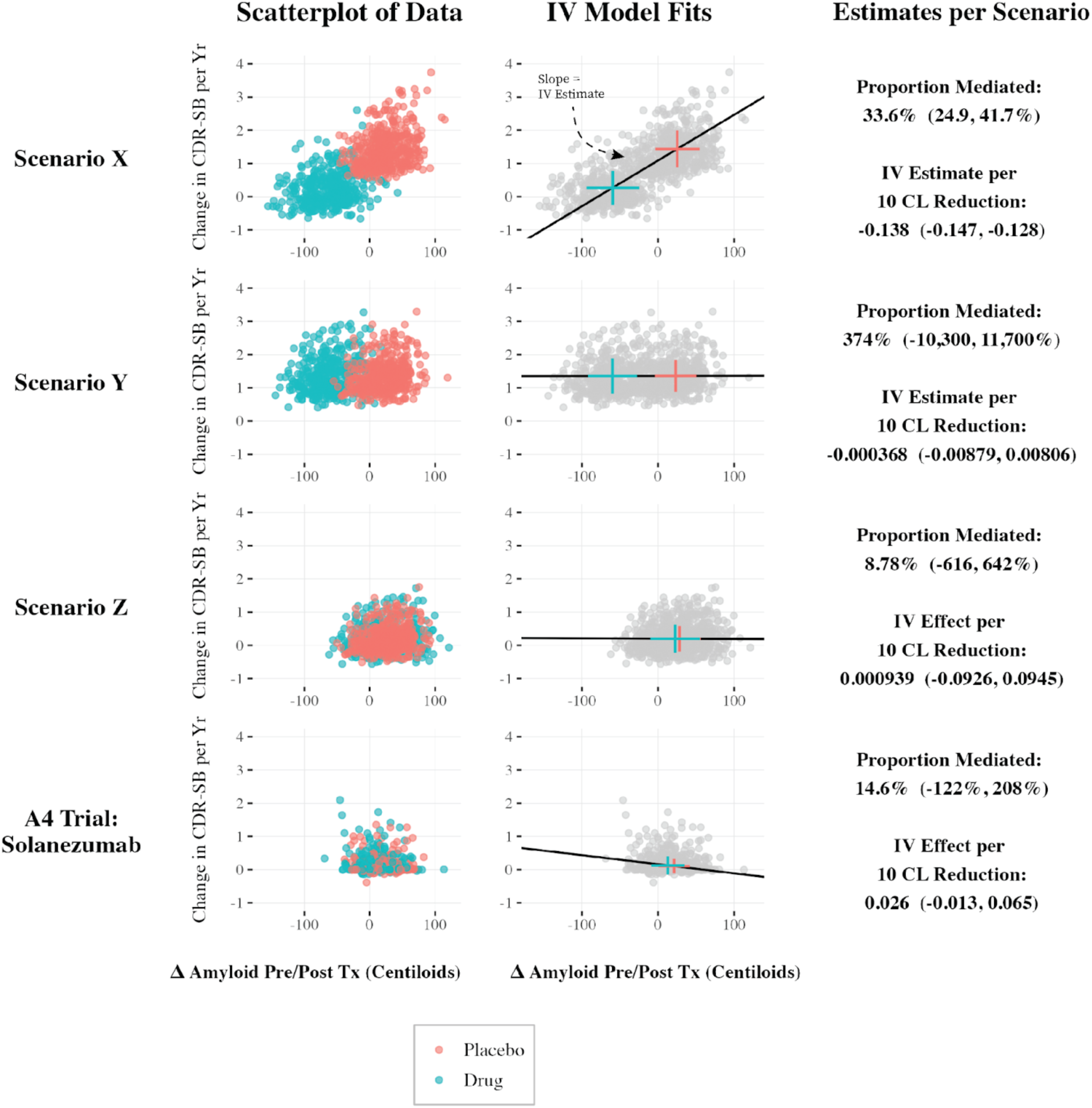
Change in cognition versus change in amyloid for simulation scenarios and the A4 trial. Rate of change in cognition is plotted against change in amyloid. Unadjusted mediation and IV estimates are presented in simulation scenarios and the A4 trial. Crosses represent +/- 1 standard deviation centered at group mean values, and the slope of regression lines connecting the intersection of crosses is equivalent to the IV estimate. Estimates of the percent mediated are not expected to provide precise results in scenarios where there is no overall effect of randomization to treatment on cognitive or functional outcome, as in scenarios Y and Z and the A4 trial. Plotted points were vertically jittered to improve visibility. IV estimates are presented as per 10 centiloid (CL) reductions and thus are sign reversed compared to the slopes of fitted lines. Scenario X: A simulated trial in which the study population experiences change in outcomes of interest. Drug X effectively removes cerebral amyloid, and benefits cognition. Scenario Y: A simulated trial in which the study population experiences change in outcomes of interest. Drug Y effectively removes amyloid, but has no impact (positive or negative) on cognition. Scenario Z: A simulated trial in which the study population does not experience meaningful change in the cognitive outcomes of interest, nor does the drug induce a change in baseline values. Drug Z does not remove amyloid, and does not impact cognition.

**Table 2.** Mediation and IV results for effects of amyloid removal on functioning and cognition in the A4 trial. Mediation results give the proportion of total effect on cognition mediated by amyloid. IV results give the effect of randomization to treatment on annual cognitive change scaled per 10 Centiloid reduction in amyloid. Cognitive outcomes include the CDR-SB, MMSE, DSST, ADCS-ADL Partner, and overall PACC scores. Adjustment set 1 includes baseline age, sex, and APOE-ε4 carrier status. Adjustment set 2 additionally includes baseline amyloid and baseline functional or cognitive measure. *Abbreviations: CDR-SB: clinical dementia rating sum of box score; MMSE: Mini-Mental State Examination; DSST: Digit Symbol Substitution Test (Wechsler Adult Intelligence Scale); ADCS-ADL: Alzheimer’s Disease Cooperative Study–Activities of Daily Living; PACC: Preclinical Alzheimer’s Cognitive Composite.*

| Method | Measure | Reported Value | Unadjusted<br>(95% CI) | Adj. Set 1<br>(95% CI) | Adj. Set 2<br>(95% CI) |
| --- | --- | --- | --- | --- | --- |
| Mediation | CDR-SB | Proportion Mediated | 0.359 ( -2.874 ,<br>3.706 ) | 0.327 ( -3.605 ,<br>3.853 ) | 0.146 ( -1.216 ,<br>2.084 ) |
| Mediation | MMSE | Proportion Mediated | 0.391 ( -9.558 ,<br>9.035 ) | 0.337 ( -10.773<br>, 7.504 ) | 0.240 ( -6.116 ,<br>7.625 ) |
| Mediation | DSST | Proportion Mediated | -0.074 ( -4.929 ,<br>8.377 ) | -0.022 ( -4.884 ,<br>8.516 ) | -0.014 ( -4.117 ,<br>2.734 ) |
| Mediation | ACDS-ADL<br>Partner<br>Score | Proportion Mediated | 0.374 ( -2.869 ,<br>4.246 ) | 0.342 ( -3.425 ,<br>3.771 ) | 0.235 ( -2.742 ,<br>2.186 ) |
| Mediation | PACC | Proportion Mediated | 0.535 ( -6.677 ,<br>11.915 ) | 0.499 ( -7.010 ,<br>9.179 ) | 0.338 ( -6.253 ,<br>6.403 ) |
| IV | CDR-SB | Effect of randomization on<br>annual rate of cognitive change,<br>scaled per 10 Cent. reduction | 0.027 ( -0.014 ,<br>0.068 ) | 0.026 ( -0.014 ,<br>0.066 ) | 0.026 ( -0.013 ,<br>0.065 ) |
| IV | MMSE | Effect of randomization on<br>annual rate of cognitive change,<br>scaled per 10 Cent. reduction | -0.012 ( -0.076 ,<br>0.052 ) | -0.010 ( -0.073 ,<br>0.052 ) | -0.009 ( -0.070 ,<br>0.051 ) |
| IV | DSST | Effect of randomization on<br>annual rate of cognitive change,<br>scaled per 10 Cent. reduction | 0.010 ( -0.213 ,<br>0.232 ) | 0.014 ( -0.204 ,<br>0.231 ) | 0.009 ( -0.203 ,<br>0.220 ) |
| IV | ACDS-ADL<br>Partner<br>Score | Effect of randomization on<br>annual rate of cognitive change,<br>scaled per 10 Cent. reduction | -0.125 ( -0.298 ,<br>0.048 ) | -0.121 ( -0.291 ,<br>0.049 ) | -0.119 ( -0.283 ,<br>0.045 ) |
| IV | PACC | Effect of randomization on<br>annual rate of cognitive change,<br>scaled per 10 Cent. reduction | -0.040 ( -0.178 ,<br>0.099 ) | -0.035 ( -0.168 ,<br>0.098 ) | -0.046 ( -0.171 ,<br>0.079 ) |

## Discussion

We reanalyzed individual-level A4 study data to evaluate whether amyloid reduction is a valid surrogate for cognitive and functional decline. We could not precisely estimate the percent of the effect of solanezumab treatment mediated by amyloid reduction, with both 0% and 100% mediation falling within the 95% confidence interval. The IV estimates, which give the effect of treatment per 10 centiloid reduction, are close to zero. This result is consistent with a previously published intent-to-treat estimate (22) and more precise than previous IV estimates in aggregated data (8). Regardless of the proportion mediated, amyloid reduction is unlikely to impact cognition over five years in populations where progression to cognitive decline is rare.

The importance of this analysis is its demonstration of an approach to evaluating amyloid reduction as a surrogate outcome – an approach that is immediately feasible and actionable with individual-level data already collected in prior trials. From mediation and IV results using the A4 trial, we cannot conclude amyloid change is a reliable surrogate for cognitive change. This is because 1) we cannot conclude amyloid largely or entirely mediates the effect of treatment on cognition and function 2) the estimated effect of a 10 Centiloid reduction on cognitive and functional change is precise and close to zero.

However, we also cannot rule out amyloid change as a reliable surrogate for change in cognition using the A4 trial data. The mediation results were inconclusive because the majority of participants did not decline and the effect of solanezumab on amyloid was small—inadequate to fully prevent treated individuals from continuing to accumulate amyloid. This is in contrast to trials of newer and newly approved drugs, lecanemab and donanemab. These drugs not only prevent amyloid accumulation, but dramatically reduce baseline amyloid levels in populations with significant cognitive decline (20,35). The IV estimates in A4 participants were close to zero since, despite the five-year follow up, the vast majority of individuals did not have any observable decline.

Rigorous methodological approaches are needed to evaluate efficacy and safety, resolve ongoing debates in the field (4–7), and inform future drug approval processes. Evaluating the relationship between amyloid reduction and cognitive and functional changes using these tools would provide clear evidence for or against using amyloid as a surrogate. If amyloid removal contributes to cognitive and functional benefits, this does not necessarily preclude a *surrogate paradox* wherein treatment causes harm, but nonetheless provides modest support for the use of amyloid as a surrogate marker for treatment efficacy (17). However, if amyloid is not the primary determinant of cognitive and functional outcomes, granting accelerated approval solely on the basis of amyloid reduction would not be justified.

IV methods offer clear benefits and preserve the advantage of randomization. Some current methodological approaches examine within-treatment-arm correlations between putative surrogate and outcome. However, if unmeasured factors influence both the surrogate and outcome, the within-arm relationship is biased for the true causal relationship between the putative surrogate and the outcome. For example, APOE-ε4 carriers may have higher levels of amyloid and thus drug treatment could remove more total amyloid. However, APOE-ε4 carriers might be expected to have the most cognitive decline due to other, preexisting biomarker changes downstream in the cascade. In this example, within a treatment arm of a drug that effectively removes amyloid, it would not be implausible to see greater cognitive decline in individuals with larger amounts of amyloid removed. In addition, noise in cognitive measures likely largely obfuscates any expected associations or inverse associations between amyloid and cognition. Thus, if we rely solely on methods that do not model differences between treatment arms, we may underestimate the utility of a potential surrogate. IV has the additional benefit of producing estimates free of regression dilution bias in contexts with measurement error in the mediator. Centiloids are measured with error (38), and thus the IV estimate produces an unattenuated estimate of the effect of amyloid on cognition. In contrast, within-arm correlations that do not preserve the benefits of randomization would be attenuated even in the absence of confounding by APOE-ε4 or other biasing factors.

The use of individual-level data confers additional advantages. First, while IV analysis is possible using aggregated data, mediation analysis is not. As shown in Figure 2, mediation analyses aid in the interpretation of IV analyses. Without mediation, it is not possible to first evaluate whether a putative surrogate fully mediates the association between treatment and the outcome (17). Estimating the percent mediated by the surrogate aids in determining whether the IV estimate is likely a correct estimate, an overestimate, or underestimate of the true causal effect. To interpret the IV estimate as the causal effect of the surrogate on the outcome (8–10) we must assume the surrogate mediates 100% of the effect of randomization; with aggregated data, this is difficult to support empirically. Second, individual-level data allow testing for non-linearity in the effect of the putative surrogate on the outcome, which has important implications for treatment optimization. Third, individual-level data allows for the assessment of effect heterogeneity in the effect of the putative surrogate on the outcome. Fourth, the IV estimator used with aggregated data is unbiased, but loses precision when the change in surrogate overlaps considerably within groups (8). More precise IV results are possible with individual-level data. Finally, individual-level adjustment may improve precision of IV estimates. In the context of drugs that more effectively remove amyloid, such improvements in precision could increase the chances of correctly identifying a valid surrogate.

While IV analysis is not common in Alzheimer’s disease research, the core ideas are a century old and have been both formally (9,10,15) and informally adopted in the context of amyloid-targeting drugs. At the 2022 Alzheimer’s Association International Conference, during a well-attended and controversial discussion on newly approved amyloid-targeting drugs, Yaning Wang presented a plot showing the mean change in cognition versus the mean change in amyloid across multiple trials (10,15,39). Examining the relationship between amyloid change and cognitive change by treatment arm or trial is both an intuitive way to visualize patterns across trials and a fundamental component of IV analyses.

Yet there are scientific risks associated with adopting IV methods in an informal manner, including: overlooking key assumptions, failing to quantify uncertainty, and lacking tools to perform formal evaluation of mechanistic effects of interest. For example, Wang suggested that the visualization he presented revealed a nonlinear relationship, arguing that greater amyloid removal led to proportionally larger cognitive benefits. While this is plausible if residual amyloid continues to drive the amyloid cascade, it was not tested statistically and no measures of uncertainty were presented. A similar plot published by a Biogen research team, for example, did not visually suggest the same nonlinear trend (10). Furthermore, arguments via visualizations of aggregated data risk obscuring important mechanistic non-linear relationships between amyloid and cognition because they only show the means across studies. There are formal statistical estimators that can test such claims of non-linearity: there is no need to “eyeball” patterns. Lastly, trial effects vary for reasons independent of amyloid removal. In A4, solanezumab did not show benefit because the placebo group did not clinically progress—results for cognitive outcomes likely would have been null even with a drug that more effectively removed amyloid. Given strong financial incentives to interpret ambiguous results in a particular direction, and entrenched scientific camps, rigorous statistical tools rather than informal interpretations are needed.

This analysis has the following limitations: First, mediation results are imprecise because the majority of study participants did not decline and solanezumab’s effects on amyloid removal were limited. As a result, we are unable to draw definitive conclusions regarding amyloid’s adequacy as a surrogate based on the A4 data alone. Relatedly, the lack of clinical progression led to estimates of precisely no cognitive benefit per amyloid reduction that may not generalize to other populations.

Second, proposed analyses are not comprehensive since other frameworks evaluate surrogacy (32,40) and in a wider range of settings (e.g., for time-to-event outcomes (17) and in meta-research (41)). Another group of authors introduced a framework for evaluating amyloid surrogate outcomes, including in individual-level data of gantenerumab (12). However, as the authors state, their methods are a tool for subgroup analysis that does not preserve the benefits of randomization for causal inference. Evaluations of safety and overall efficacy, examining within-treatment-arm correlations, and other approaches (18,19), are still valuable in this context; these approaches should be viewed as complementary to ours because the various approaches require different assumptions. Nonetheless, mediation (19) and IV analyses are unique in that they can adjust for within arm confounding and preserve the benefits of randomization. They are valuable as methodological elements of a more complete framework to evaluate amyloid removal as a surrogate outcome for cognition.

In conclusion, using mediation and IV methods to analyze individual-level data from trials of amyloid-targeting drugs can improve our understanding of amyloid as a surrogate outcome for cognition (or for other patient-centered outcomes). In this instance, mediation results were imprecise because progression was rare and because solanezumab did not effectively remove amyloid. IV estimates were close to zero due to the lack of clinical progression. A logical next step is to perform these analyses in trial populations with clinical progression using interventions that effectively remove amyloid.

## Data Availability

Data were obtained from the Anti-Amyloid Treatment in Asymptomatic Alzheimer's Disease (A4) and Longitudinal Evaluation of Amyloid Risk and Neurodegeneration (LEARN) Study Data Package. Data is available to qualified researchers with a short application at a4studydata.org.

https://www.a4studydata.org/

## Acknowledgments

The A4 Study was a secondary prevention trial in preclinical Alzheimer’s disease, aiming to slow cognitive decline associated with brain amyloid accumulation in clinically normal older individuals. The A4 Study was funded by a public-private-philanthropic partnership, including funding from the National Institutes of Health-National Institute on Aging, Eli Lilly and Company, Alzheimer’s Association, Accelerating Medicines Partnership, GHR Foundation, an anonymous foundation, and additional private donors, with in-kind support from Avid Radiopharmaceuticals, Cogstate, Albert Einstein College of Medicine, and the Foundation for Neurologic Diseases. The companion observational Longitudinal Evaluation of Amyloid Risk and Neurodegeneration (LEARN) Study was funded by the Alzheimer’s Association and GHR Foundation. The A4 and LEARN Studies were led by Dr. Reisa Sperling at Brigham and Women’s Hospital, Harvard Medical School, and Dr. Paul Aisen at the Alzheimer’s Therapeutic Research Institute (ATRI) at the University of Southern California. The A4 and LEARN Studies were coordinated by ATRI at the University of Southern California, and the data are made available under the auspices of Alzheimer’s Clinical Trial Consortium through the Global Research & Imaging Platform (GRIP). The complete A4 Study Team list is available on: https://www.actcinfo.org/a4-study-team-lists/ We would like to acknowledge the dedication of the study participants and their study partners who made the A4 and LEARN Studies possible.

## Footnotes

1 The IV literature typically refers to what we call the mediator as the exposure. We avoid this standard terminology to help illustrate the connection between mediation and IV and maintain consistency in our use in terminology. Hence, for both types of analyses, we refer to amyloid change as the mediator or the surrogate outcome.

## References

1. Alexander GC, Knopman DS, Emerson SS, Ovbiagele B, Kryscio RJ, Perlmutter JS, et al. Revisiting FDA Approval of Aducanumab. N Engl J Med. 2021 Aug 26;385(9):769–71.

2. Knopman DS, Hershey L. Implications of the Approval of Lecanemab for Alzheimer Disease Patient Care. Neurology. 2023 Oct 3;101(14):610–20.

3 Rosen J, Jessen F. Patient eligibility for amyloid-targeting immunotherapies in Alzheimer’s disease. The Journal of Prevention of Alzheimer’s Disease. 2025 Apr 1;12(4):100102.

4 Aisen P, Bateman RJ, Crowther D, Cummings J, Dwyer J, Iwatsubo T, et al. The case for regulatory approval of amyloid-lowering immunotherapies in Alzheimer’s disease based on clearcut biomarker evidence. Alzheimer’s & Dementia. 2025;21(1):e14342.

5. Selkoe DJ. Treatments for Alzheimer’s disease emerge. Science. 2021 Aug 6;373(6555):624–6.

6. Planche V, Schindler S, Knopman DS, Frisoni G, Galasko D, Grill JD, et al. The science does not yet support regulatory approval of amyloid-targeting therapies for Alzheimer’s disease based solely on biomarker evidence. Alzheimer’s & Dementia. 2025;21(4):e70068.

7 Thambisetty M, Howard R, Glymour MM, Schneider LS. Alzheimer’s drugs: Does reducing amyloid work? Science. 2021 Oct 29;374(6567):544–5.

8. Ackley SF, Zimmerman SC, Brenowitz WD, Tchetgen EJT, Gold AL, Manly JJ, et al. Effect of reductions in amyloid levels on cognitive change in randomized trials: instrumental variable meta-analysis. BMJ. 2021 Feb 25;372:n156.

9 Ackley SF, Wang J, Chen R, Power MC, Allen IE, Glymour MM. Estimated effects of amyloid reduction on cognitive change: A Bayesian update across a range of priors. Alzheimer’s & Dementia. 2024;20(2):1149–55.

10. Pang M, Zhu L, Gabelle A, Gafson AR, Platt RW, Galvin JE, et al. Effect of reduction in brain amyloid levels on change in cognitive and functional decline in randomized clinical trials: An instrumental variable meta-analysis. Alzheimers Dement. 2022 Aug 31;

11 Ren S, Singh J, Gsteiger S, Cogley C, Reed B, Abrams KR, et al. Evaluating amyloid-beta as a surrogate endpoint in trials of anti-amyloid drugs in Alzheimer’s disease: a Bayesian meta-analysis [Internet]. arXiv; 2025 [cited 2025 Apr 25]. Available from: http://arxiv.org/abs/2504.06807

12. Wang G, Li Y, Xiong C, Benzinger TLS, Gordon BA, Hassenstab J, et al. Examining amyloid reduction as a surrogate endpoint through latent class analysis using clinical trial data for dominantly inherited Alzheimer’s disease. Alzheimers Dement. 2024 Feb 23;20(4):2698–706.

13. Chen T, Hutchison RM, Rubel C, Murphy J, Xie J, Montenigro P, et al. A Statistical Framework for Assessing the Relationship between Biomarkers and Clinical Endpoints in Alzheimer’s Disease. The Journal of Prevention of Alzheimer’s Disease. 2024 Oct 1;11(5):1228–40.

14. Ackley SF, Glymour MM. Comment on “Effect of reduction in brain amyloid levels on change in cognitive and functional decline in randomized clinical trials: An updated instrumental variable meta-analysis.” Alzheimer’s & Dementia. 2023;19(3):1099–100.

15. Ackley SF, Elahi F, Glymour MM. Instrumental variable meta-analysis of aggregated randomized drug trial data for evaluating proposed target mechanisms. BMJ. 2021 Feb 25;372:n346.

16. VanderWeele TJ. Mediation Analysis: A Practitioner’s Guide. Annual Review of Public Health. 2016 Mar 18;37(Volume 37, 2016):17–32.

17. Elliott MR. Surrogate Endpoints in Clinical Trials. Annual Review of Statistics and Its Application. 2023 Mar 9;10(Volume 10, 2023):75–96.

18. Baker SG. Five criteria for using a surrogate endpoint to predict treatment effect based on data from multiple previous trials. Stat Med. 2018 Feb 20;37(4):507–18.

19. Christensen R, Ciani O, Manyara AM, Taylor RS. Surrogate endpoints: a key concept in clinical epidemiology. Journal of Clinical Epidemiology [Internet]. 2024 Mar 1 [cited 2025 Apr 24];167. Available from: https://www.jclinepi.com/article/S0895-4356(23)00340-2/fulltext

20. Sims JR, Zimmer JA, Evans CD, Lu M, Ardayfio P, Sparks J, et al. Donanemab in early symptomatic Alzheimer disease: the TRAILBLAZER-ALZ 2 randomized clinical trial. JAMA. 2023;

21. Sperling RA, Rentz DM, Johnson KA, Karlawish J, Donohue M, Salmon DP, et al. The A4 Study: Stopping AD Before Symptoms Begin? Science Translational Medicine. 2014 Mar 19;6(228):228fs13–228fs13.

22. Insel PS, Donohue MC, Sperling R, Hansson O, Mattsson-Carlgren N. The A4 study: β-amyloid and cognition in 4432 cognitively unimpaired adults. Annals of Clinical and Translational Neurology. 2020;7(5):776–85.

23. Grober E, Lipton RB, Sperling RA, Papp KV, Johnson KA, Rentz DM, et al. Associations of Stages of Objective Memory Impairment With Amyloid PET and Structural MRI. Neurology. 2022 Mar 29;98(13):e1327–36.

24. Amariglio RE, Grill JD, Rentz DM, Marshall GA, Donohue MC, Liu A, et al. Longitudinal Trajectories of the Cognitive Function Index in the A4 Study. J Prev Alzheimers Dis. 2024 Aug 1;11(4):838–45.

25. Sperling RA, Donohue MC, Raman R, Rafii MS, Johnson K, Masters CL, et al. Trial of Solanezumab in Preclinical Alzheimer’s Disease. New England Journal of Medicine. 2023 Sep 20;389(12):1096–107.

26. Papp KV, Rentz DM, Maruff P, Sun CK, Raman R, Donohue MC, et al. The Computerized Cognitive Composite (C3) in A4, an Alzheimer’s Disease Secondary Prevention Trial. J Prev Alzheimers Dis. 2021 Jan 1;8(1):59–67.

27. Glymour MM, Weuve J, Berkman LF, Kawachi I, Robins JM. When Is Baseline Adjustment Useful in Analyses of Change? An Example with Education and Cognitive Change. American Journal of Epidemiology. 2005 Aug 1;162(3):267–78.

28. Imai K, Keele L, Tingley D. A general approach to causal mediation analysis. Psychological Methods. 2010;15(4):309–34.

29. MacKinnon DP, Fairchild AJ, Fritz MS. Mediation Analysis. Annual Review of Psychology. 2007 Jan 1;58(Volume 58, 2007):593–614.

30. Richiardi L, Bellocco R, Zugna D. Mediation analysis in epidemiology: methods, interpretation and bias. International Journal of Epidemiology. 2013 Oct 1;42(5):1511–9.

31. Baron RM, Kenny DA. The moderator–mediator variable distinction in social psychological research: Conceptual, strategic, and statistical considerations. Journal of Personality and Social Psychology. 1986;51(6):1173–82.

32. Prentice RL. Surrogate endpoints in clinical trials: definition and operational criteria. Stat Med. 1989 Apr;8(4):431–40.

33. Stock JH, Trebbi F. Retrospectives: Who Invented Instrumental Variable Regression? Journal of Economic Perspectives. 2003 Sep;17(3):177–94.

34. Meleth S. Adherence Adjusted Estimates in Randomized Clinical Trials. In: Piantadosi S, Meinert CL, editors. Principles and Practice of Clinical Trials [Internet]. Cham: Springer International Publishing; 2019 [cited 2025 Apr 23]. p. 1–18. Available from: 10.1007/978-3-319-52677-5_124-1

35. van Dyck CH, Swanson CJ, Aisen P, Bateman RJ, Chen C, Gee M, et al. Lecanemab in Early Alzheimer’s Disease. New England Journal of Medicine. 2023 Jan 5;388(1):9–21.

36. Imai K, Keele L, Tingley D, Yamamoto T. Causal Mediation Analysis Using R. In: Vinod HD, editor. Advances in Social Science Research Using R. New York, NY: Springer; 2010. p. 129–54.

37. Li Y, Albert JM. Semi-parametric model approach to causal mediation analysis for longitudinal data. Statistical Modelling. 2025 Feb 7;1471082X241306911.

38. Collij LE, Bollack A, La Joie R, Shekari M, Bullich S, Roé-Vellvé N, et al. Centiloid recommendations for clinical context-of-use from the AMYPAD consortium. Alzheimers Dement. 2024 Dec;20(12):9037–48.

39. Could Benefit of Plaque Removal Grow in Time? | ALZFORUM [Internet]. [cited 2025 May 6]. Available from: https://www.alzforum.org/news/conference-coverage/could-benefit-plaque-removal-grow-time

40. Ciani O, Manyara AM, Davies P, Stewart D, Weir CJ, Young AE, et al. A framework for the definition and interpretation of the use of surrogate endpoints in interventional trials. eClinicalMedicine [Internet]. 2023 Nov 1 [cited 2025 May 16];65. Available from: https://www.thelancet.com/journals/eclinm/article/PIIS2589-5370(23)00460-1/fulltext

41. Ciani O, Buyse M, Garside R, Pavey T, Stein K, Sterne JAC, et al. Comparison of treatment effect sizes associated with surrogate and final patient relevant outcomes in randomised controlled trials: meta-epidemiological study. BMJ. 2013 Jan 29;346:f457.

42. Muir RT, Hill MD, Black SE, Smith EE. Minimal clinically important difference in Alzheimer’s disease: Rapid review. Alzheimers Dement. 2024 May;20(5):3352–63.

