## Supplemental Material for "Evaluation of Amyloid Removal as a Surrogate for Cognitive Decline: Pilot Analysis in Individual-Level Data from the A4 Study of Solanezumab"

### Supplemental Tables

**Table S1:** Cognitive outcomes assessed. Descriptions of corresponding cognitive domains, scale ranges, and established cutoffs for mild cognitive impairment (MCI) or dementia are included.

Abbreviations: *CDR-SB: clinical dementia rating sum of box score; MMSE: Mini-Mental State Examination; DSST: Digit Symbol Substitution Test (Wechsler Adult Intelligence Scale); ADCS-ADL: Alzheimer's Disease Cooperative Study–Activities of Daily Living.*

| **Cognitive Outcome** | **Cognitive Domain** | **Scale Range** | **MCI/Dementia Cutoff** |
|---|---|---|---|
| CDR-SB | Global cognition & function | 0-18 (higher = worse) | >=0.5 indicative of impairment |
| MMSE | Global cognition | 0-30 (higher = better) | <24 suggests MCI, <20 suggests dementia |
| DSST | Processing speed & executive function | 0-133 (higher = better) | No established cutoff |
| ADCS-ADL | Functional abilities | 0-78 (higher = better) | Lower scores indicate impairment, but no specific cutoff |
| PACC | Episodic memory, executive function, global cognition | Composite z-score (higher = better) | No fixed cutoff; used to detect early decline |

**Table S2.** A version of Table 1 showing pre-/post-treatment differences in cognitive measures rather than modeled annual change.

| | Placebo (N=414) | Solanezumab (N=391) | Overall (N=805) |
|---|---|---|---|
| **Age** | 71.48 (4.47) | 71.47 (4.43) | 71.48 (4.45) |
| **Sex** | | | |
| Female | 245 (59.2%) | 229 (58.6%) | 474 (58.9%) |
| Male | 169 (40.8%) | 162 (41.4%) | 331 (41.1%) |
| **APOE4 Allele Copies** | | | |
| 0 | 159 (38.4%) | 149 (38.1%) | 308 (38.3%) |
| 1 | 222 (53.6%) | 207 (52.9%) | 429 (53.3%) |
| 2 | 33 (8.0%) | 35 (9.0%) | 68 (8.4%) |
| **Baseline Amyloid Burden on PET, centiloids** | 65.92 (31.08) | 65.17 (30.85) | 65.56 (30.95) |
| **Pre/Post Change in Amyloid PET, centiloids** | 21.14 (20.66) | 12.77 (21.54) | 17.07 (21.49) |
| **Baseline CDR-SB Score** | 0.05 (0.17) | 0.05 (0.16) | 0.05 (0.17) |
| **Change in CDR-SB Score pre/post RCT** | 0.57 (1.18) | 0.64 (1.36) | 0.60 (1.27) |
| **Baseline MMSE Score** | 28.79 (1.24) | 28.86 (1.20) | 28.82 (1.22) |
| **Change in MMSE Score pre/post RCT** | -0.39 (2.04) | -0.43 (2.25) | -0.41 (2.14) |
| **Baseline DSST Score** | 49.35 (9.63) | 49.17 (10.22) | 49.26 (9.92) |
| **Change in DSST Score pre/post RCT** | -2.46 (7.68) | -2.49 (7.35) | -2.47 (7.52) |
| **Baseline ADL Partner Score** | 43.47 (2.67) | 43.56 (2.39) | 43.51 (2.53) |
| **Change in ADL Partner Score pre/post RCT** | -1.57 (5.33) | -2.07 (5.59) | -1.81 (5.46) |
| **Baseline PACC Score** | 0.10 (2.60) | 0.24 (2.69) | 0.17 (2.64) |
| **Change in PACC Score pre/post RCT** | -1.13 (4.14) | -1.24 (4.39) | -1.18 (4.26) |

**Table S3.** Sensitivity analysis. Results of mediation and IV analyses but here using individual-level pre-/post-treatment differences in functional and cognitive scores instead of individual-level modeled annual change in cognition.

| Method | Measure | Reported Value | Unadjusted (95% CI) | Adjustment Set 1 (95% CI) | Adjustment Set 2 (95% CI) |
|---|---|---|---|---|---|
| Mediation | CDR-SB | Proportion Mediated | 0.260 ( -5.625 , 4.439 ) | 0.215 ( -3.337 , 4.815 ) | 0.076 ( -1.600 , 2.604 ) |
| Mediation | MMSE | Proportion Mediated | 0.272 ( -9.449 , 6.056 ) | 0.266 ( -6.396 , 5.575 ) | 0.127 ( -5.858 , 7.747 ) |
| Mediation | DSST | Proportion Mediated | 0.079 ( -6.696 , 6.277 ) | 0.107 ( -6.028 , 4.147 ) | 0.052 ( -3.434 , 4.244 ) |
| Mediation | ADL Partner Score | Proportion Mediated | 0.356 ( -3.827 , 3.902 ) | 0.328 ( -3.030 , 3.918 ) | 0.252 ( -2.155 , 2.805 ) |
| Mediation | PACC | Proportion Mediated | 0.454 ( -14.501 , 10.520 ) | 0.418 ( -11.109 , 7.917 ) | 0.314 ( -5.971 , 5.969 ) |
| IV | CDR-SB | Effect of randomization on annual rate of cognitive change, scaled per 10 Cent. reduction | 0.093 ( -0.117 , 0.304 ) | 0.090 ( -0.116 , 0.295 ) | 0.093 ( -0.109 , 0.294 ) |
| IV | MMSE | Effect of randomization on annual rate of cognitive change, scaled per 10 Cent. reduction | -0.043 ( -0.397 , 0.311 ) | -0.039 ( -0.387 , 0.309 ) | -0.005 ( -0.335 , 0.325 ) |
| IV | DSST | Effect of randomization on annual rate of cognitive change, scaled per 10 Cent. reduction | -0.044 ( -1.288 , 1.199 ) | -0.034 ( -1.259 , 1.190 ) | -0.098 ( -1.283 , 1.088 ) |
| IV | ADL Partner Score | Effect of randomization on annual rate of cognitive change, scaled per 10 Cent. reduction | -0.605 ( -1.511 , 0.301 ) | -0.591 ( -1.482 , 0.300 ) | -0.578 ( -1.429 , 0.273 ) |
| IV | PACC | Effect of randomization on annual rate of cognitive change, scaled per 10 Cent. reduction | -0.133 ( -0.835 , 0.569 ) | -0.122 ( -0.800 , 0.556 ) | -0.177 ( -0.826 , 0.472 ) |

### Supplemental Figures

**Figure S1.** Group-level trajectories of functional and cognitive outcomes across study visits. The left panels display median values with interquartile ranges (IQR), while the right panels show means with ±1 standard deviation (SD) for each visit. Visit 66 corresponds to the end of the nearly 5-year study period. Overall, both median and mean trajectories indicate minimal change over time across most measures.

*Abbreviations: IQR: interquartile range; SD: standard deviation; TX: treatment.*

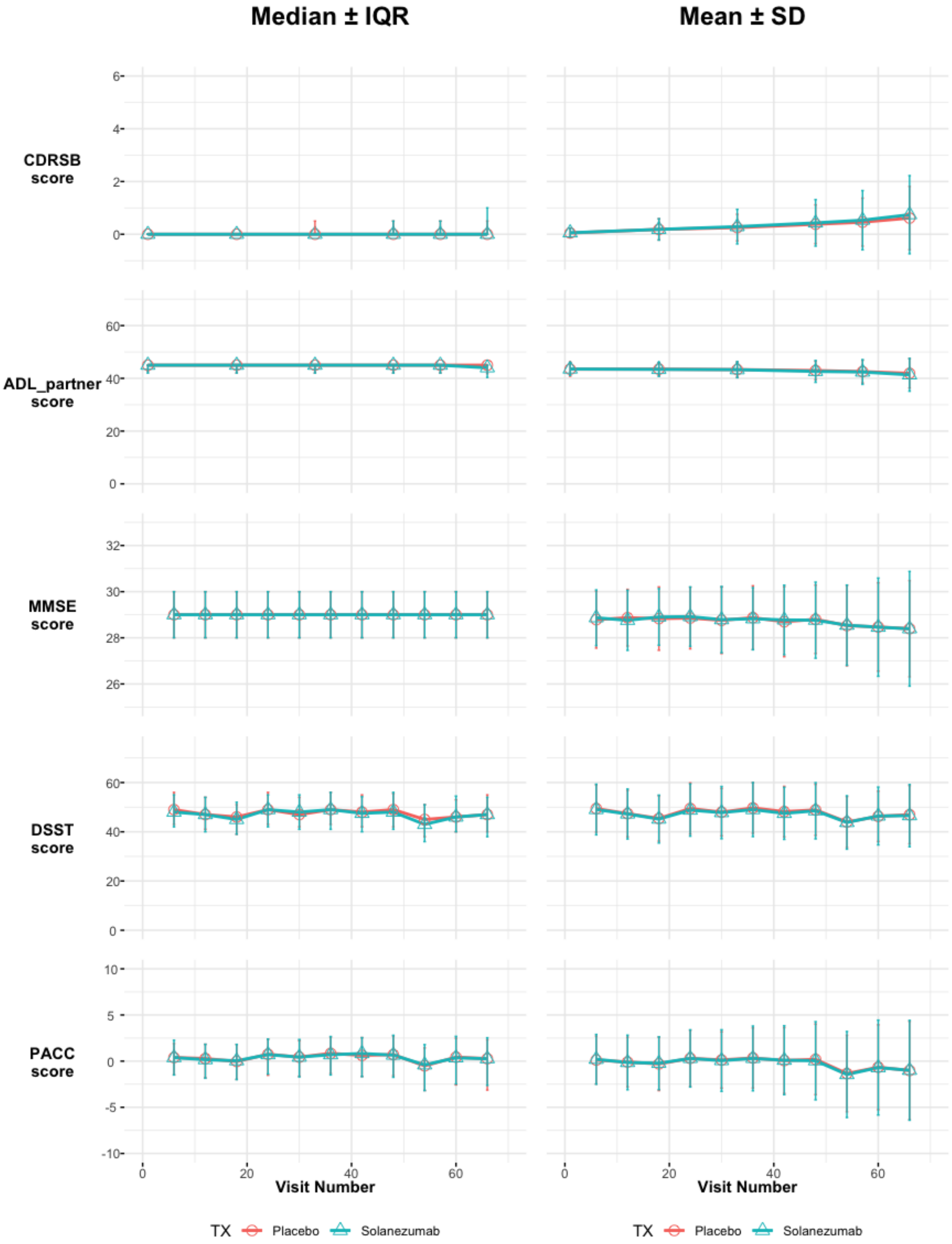

Median ± IQR
Mean ± SD
CDRSB score
ADL_partner score
MMSE score
DSST score
PACC score
Visit Number
Visit Number
TX
Placebo
Solanezumab
TX
Placebo
Solanezumab

**Figure S2.** Comparison of the distribution of modeled annual functional and cognitive change (left) to observed pre/post-treatment cognitive differences (right). Pre/post-treatment functional and cognitive change is over approximately 5 years versus modeled annual functional and cognitive change. Modeled annual functional and cognitive change and pre/post-treatment cognitive difference are comparable and show very similar patterns. (Pink= placebo; teal = solanezumab.)

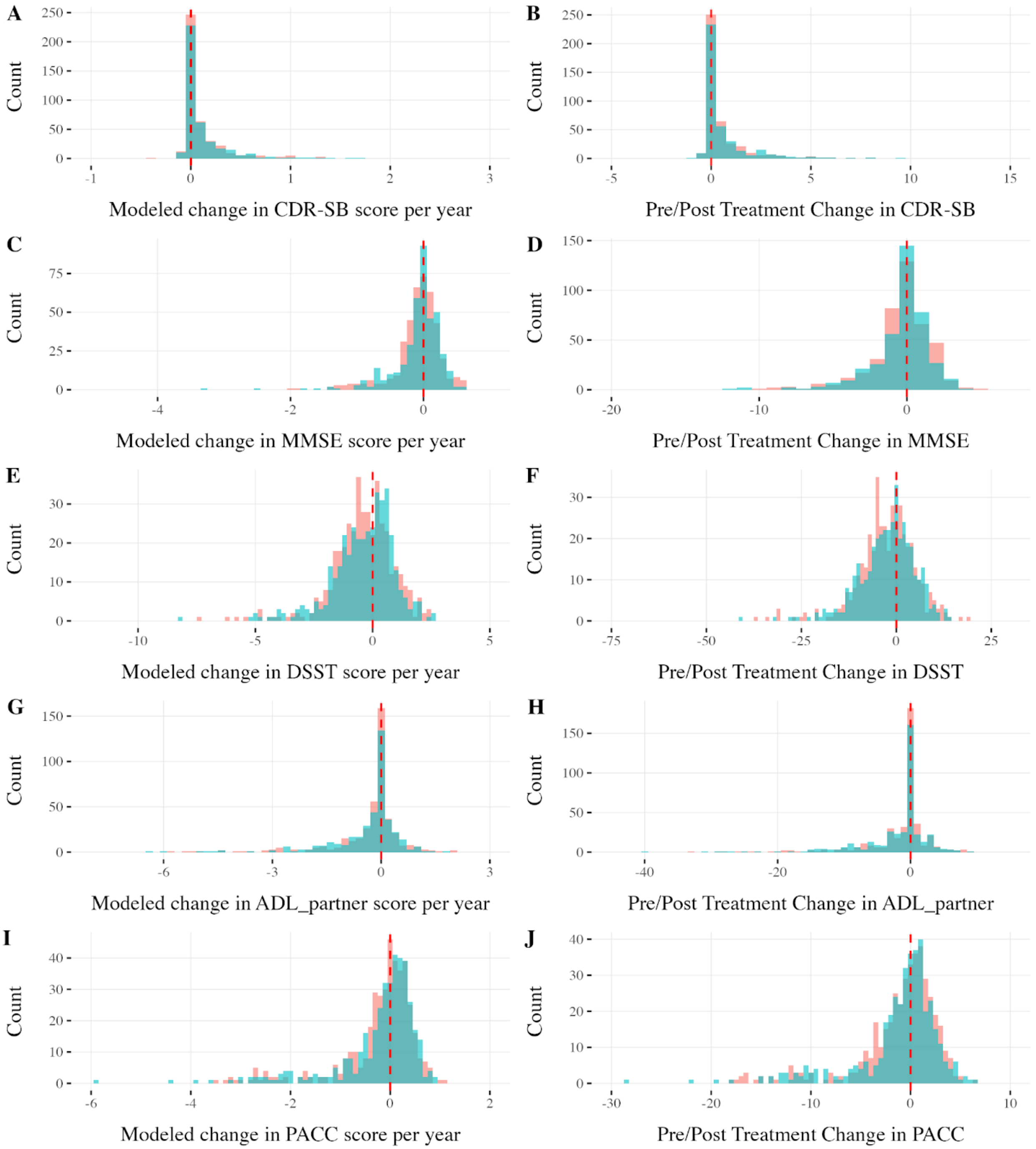
A
Count
250
200
150
100
50
0
-1
0
1
2
3
Modeled change in CDR-SB score per year
B
Count
250
200
150
100
50
0
-5
0
5
10
15
Pre/Post Treatment Change in CDR-SB
C
Count
75
50
25
0
-4
-2
0
Modeled change in MMSE score per year
D
Count
150
100
50
0
-20
-10
0
Pre/Post Treatment Change in MMSE
E
Count
30
20
10
0
-10
-5
0
5
Modeled change in DSST score per year
F
Count
30
20
10
0
-75
-50
-25
0
25
Pre/Post Treatment Change in DSST
G
Count
150
100
50
0
-6
-3
0
3
Modeled change in ADL_partner score per year
H
Count
150
100
50
0
-40
-20
0
Pre/Post Treatment Change in ADL_partner
I
Count
40
30
20
10
0
-6
-4
-2
0
2
Modeled change in PACC score per year
J
Count
40
30
20
10
0
-30
-20
-10
0
10
Pre/Post Treatment Change in PACC

**Figure S3.** Mean change in CDR-SB versus mean change in amyloid by dose escalation visit number for treated groups (black dots) and placebo group (red dots). Slope of the plotted line gives the IV estimate for the effect of amyloid change on CDR-SB change.

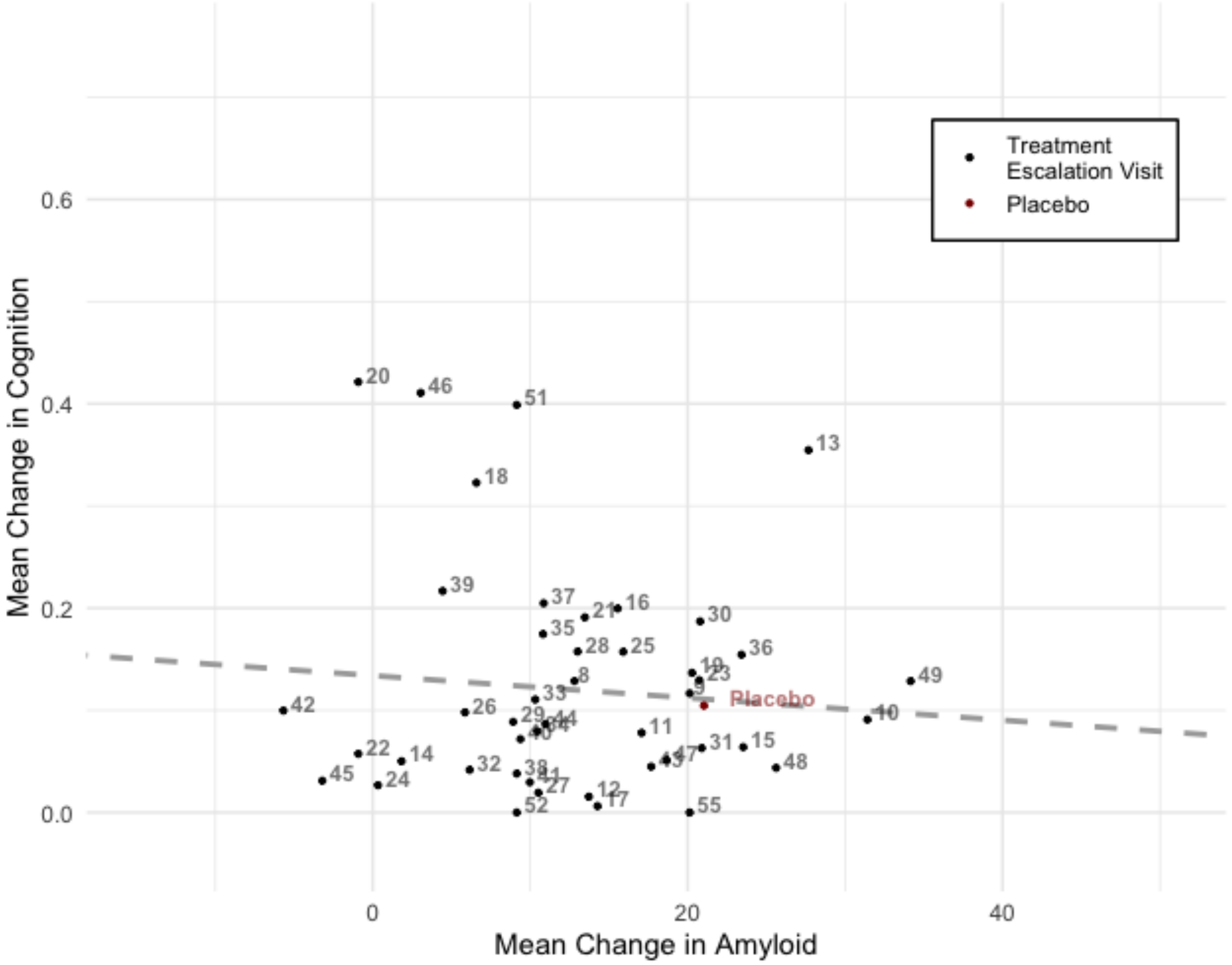
